# Parenthood, Mental Disorders, and Symptoms Through Adulthood: A Total Population Study

**DOI:** 10.1101/2024.10.29.24315908

**Authors:** ML Andersen, HF Sunde, RK Hart, FA Torvik

**Affiliations:** Centre for Fertility and Health, Norwegian Institute of Public Health, Oslo, Norway.; Department of Psychology, University of Oslo, Oslo, Norway.; Institute of Health and Society, University of Oslo, Oslo, Norway.

**Keywords:** Mental health, parenthood, childlessness, social inequality, fertility

## Abstract

During recent decades, parenthood has declined in many Western countries. Simultaneously, mental disorders have become more prevalent. We investigated the link between parenthood and mental health in the entire Norwegian-born population aged 31 to 80 from 2006 to 2019 (n=2,234,087). We used logistic regression models on national register data and included sibling- and twin-matched analyses to address unobserved confounders. Parenthood was associated with a lower risk of mental disorders, including depressive and anxiety disorders. For symptoms related to mental disorders, fathers had a reduced risk, while mothers had a slightly elevated risk. Mental health disparities between parents and non-parents were greater among men than among women and persisted across adulthood, before reducing at older ages. Our main findings were largely consistent in sibling- and twin-matched designs. The disparity between parents and non-parents increased over the study period, suggesting stronger selection into parenthood. Our findings highlight parenthood as a significant indicator of mental health inequalities, with its importance growing over time.

## 1 Introduction

Finding a partner and raising children significantly alter the daily experience and is plausibly related to mental health. Mental disorders are responsible for the most working years lost in the Norwegian population [1] and are also considered among the leading causes of disease burden worldwide [2]. Consequently, understanding the factors related to the risk of mental disorders is important. Whereas associations with psychosocial factors such as sex and educational attainment have been extensively documented [3–5], links with parenthood have so far been underexplored. Most studies on parenthood and mental health focus either on health-based selection into parenthood [6, 7], on the risk of ante- and post-natal depression [8, 9], or on the association between parenthood and mental health in small samples of limited age-range. Furthermore, over the last decades there have been considerable changes in fertility behaviour, as people are having fewer children and postponing their first birth [10]. Concurrently with the decline in fertility, there has also been a marked increase in mental distress in multiple groups [11]. Given the co-occurrence of increased mental disorders and declining fertility in many countries, a possible link between the two phenomena must be explored. As childlessness has become increasingly common, it is important that we better understand how parenthood and outcomes such as mental health are linked. Childless adults will not have the natural care-givers parents have in old age (i.e. children and grandchildren). Thus, from a healthcare policy perspective it is especially important that we understand how this group may differ in terms of their health. For this reason, it is also important to study if the association between mental health and fertility is changing over time. Furthermore, while there has been increased focus on how socioeconomic inequalities relate to mental health in the population, little focus has been directed towards how parenthood can be a marker of mental health inequalities. Fertility may be an important but less studied social indicator of mental health inequalities.

The relationship between mental health and parenthood has been examined across various disciplines, yet the results are diverging. Some studies focus on negative changes in mental health during the period following childbirth [12], which leaves long-term associations between parenthood and mental health unaddressed. A German study found that parenthood was associated with better mental health, in particular among men, when comparing those living with at least one child to childless individuals [13]. Yet, when comparing empty-nest parents to childless adults, a Norwegian study only found small positive associations with women’s well-being, and no association with men’s well-being [14]. Another Norwegian study looking at parenthood and symptoms of anxiety and depression in a large population based cohort found no differences between parents and non-parents [15]. This study also focused on a very narrow age-range (30-49), and by doing so it excluded most empty-nesters. Finally, some studies even find a negative impact of parenthood on mental health, with some types of parenthood being associated with higher levels of depression [16]. One potential explanation for these discrepancies is the lack of consistency in the methods and timing of comparisons between parents and non-parents. For example, studies also vary in how they operationalise mental health. Previous work variously investigates diagnoses of mental disorders, symptoms of anxiety and depression, or even subjective-wellbeing. Although related, such measures are qualitatively different [17], and their associations with parenthood could vary. Additionally, much of the existing literature predominantly focuses on shorter time spans, leaving long-term associations unknown.

There are few studies on the relationship between parenthood and health outcomes at older ages, although some studies indicate a lower mortality among parents [18, 19]. A multinational study incorporating data on elderly from Finland, Australia, and the Netherlands found disparate links between parenthood and measures of depression. In Finland, formerly married childless men were less likely to report feelings of depression, while in Australia, it was formerly married childless women who were less likely to report such feelings. In the Netherlands, married men were unlikely to report depression regardless of parent-status [20]. However, it is important to note that the surveys included in this study had different measures of depression. A U.S. study focused on those aged 70 or older also found no significant evidence of an increase in loneliness or depression among childless individuals [21]. Nonetheless, the authors did find that sex had a significant impact on how childlessness and marital status related to psychological wellbeing. While life course approaches are scarce among studies looking at parenthood and mental health, some indications may be found in studies of subjective wellbeing, which is related to mental health. Cetre et al. [22] found that the association between parenthood and subjective wellbeing was only positive in developed countries. Using German panel data, they also showed how raw measures of life satisfaction remained higher for parents than non-parents throughout the life course. However, previous research has found evidence of an anticipation and adaptation effect when it comes to life events and subjective wellbeing measures [23]. Consequently, subjective wellbeing is likely not a good indicator of the presence of more chronic mental disorders or symptoms at higher ages. Based on the variation in findings across past studies, one may hypothesise that the health impact of parenthood vary across the life course.

Besides the previously discussed health outcomes, parents and non-parents differ in many ways. In Norway, childless men are typically more likely to be single, have lower educational attainment, lower income, and receive more transfer payments [24]. Among women on the other hand, the highest rates of childlessness have historically been found amid the most educated [25]. However, in more recent female birth cohorts, the highest rates of childlessness are also found among the least educated [26]. Cetre et al. also find evidence of selection into parenthood in Germany based on individuals’ happiness prior to childbearing [22]. While these characteristics associated with parenthood are well documented, research on how parenthood is differently associated with outcomes such as mental health conditional on factors like socioeconomic status is scarce. As illustrated, the results of pre-existing research are mixed and make it difficult to conclude about the overall association between parenthood and mental health over the life course. There is also little research on how these associations are shaped by factors like socioeconomic status or change over time. However, the different findings across studies do suggest that factors such as age, national and political context, partnership, and gender likely influence the consequences of parenthood. In the present study, we apply a unique population-wide dataset to study the links between fertility and mental health for an entire population. We therefore avoid many of the issues connected to smaller sample sizes and self-reported measures of health seen in the majority of previous work on this topic. With access to primary health care data for an entire population we also have the opportunity to study how associations between fertility and mental health may vary across subgroups such as age, gender, and socioeconomic status. As far as we are aware, this is the only primary care dataset that covers an entire nation. An advantage is that these data include symptoms of mental disorders, not only diagnoses. When an association between parenthood and mental health is seen, it can arise for several reasons: **1)** having children may improve mental health, **2)** individuals with good mental health may be more likely to have children, or **3)** common factors may influence both mental health and the likelihood of becoming a parent. By comparing same-sex siblings and monozygotic twin pairs we can account for the genetic and environmental influences shared in families, thus ruling out shared causes. In this way, this study fills some of the gaps in the current literature by studying both the overall life course associations as well as possible interactions with factors like socioeconomic status.

Using register data covering the entire population of Norway, we investigate how parenthood is related to mental health by addressing the following four research questions:

**RQ1:** How is parenthood related to mental health among men and women across the life-course, after adjustment for observed confounders, and within sibships?

**RQ2:** Do differences in mental health between parents and non-parents vary by socioeconomic background, partnership status, and age at first birth?

**RQ3:** Has the relationship between mental health and parenthood changed over the past 15 years?

**RQ4:** How is parenthood associated with different diagnoses and kinds of symptoms?

## 2 Methods

### Sample

Our study population is based on the Norwegian Central Population Register. This register includes anyone who has been registered as residing in Norway since the 1960 census and contains information about time of birth, death, and parent identifiers until the end of 2020. This was linked to educational and socioeconomic register data and diagnostic data from medical records covering the years 2006-2019. The data from these different sources were joined using pseudonymised individual-specific personal identification numbers. To minimise incomplete data, we limited our analyses to only include Norwegian-born individuals who also were registered as residing in Norway during the observation period (2006-2019). In order for individuals to only belong to one group (i.e. parent or childless), we focus on completed fertility at age 45. Consequently, only those born in 1975 or earlier are included. This leaves in total 28,823,322 person-years stemming from 2,234,087 unique individuals in our dataset.

To adjust for potential genetic and environmental effects shared between siblings that confound the relationship between fertility and mental health, we also used parent identifiers to create a sub-sample of same-sex full-siblings in the population. When limiting the sample to only those with at least one full same-sex sibling, we have 13,036,358 observations based on 434,070 unique sets of same-sex siblings (202,613 female sets and 231,457 male sets). To achieve full adjustment for genetic factors, we also identified 2,132 female and 2,380 male monozygotic twin pairs from the Norwegian Twin Registry.

### Measures

#### Parenthood and age at first birth

Data from the Norwegian Central Population Register is used to identify all first births in the population. We defined parents as individuals who were registered with a first birth before or at age 45, while we defined those registered without any births by age 45 as childless. We also use the Norwegian Central Population Register to identify the individual’s age at first live birth. Women are considered to have their first child at an “early” age if they had a first birth before 25. Conversely, if they had their first birth at age 33 or later, we consider them as of “advanced” age at first birth. Men are grouped in a similar way, with cut offs at 27 and 35 years. We also calculate the number of children at age 45.

#### Mental disorders and symptoms

Information about primary health care utilisation comes from the Norwegian Control and Payment of Health Reimbursements Database (KUHR), which was established in 2006. The KUHR database contains information from different contact points within the health care system, with the majority of the data coming from general practitioners. When practitioners submit their reimbursement claims to the Norwegian Health Economics Administration (Helfo), they use the International Classification of Primary Care (ICPC-2) system to register the reason for a patient’s visit [27]. Codes P01-P29 are used to indicate psychological symptoms, while codes P70-P99 refer to diagnoses of mental disorders. As general practitioners are required to submit these forms to receive reimbursements, it is unlikely that visits are under-reported in our data. However, the oldest age groups in our dataset are likely to have fewer reported visits than expected. This is because health care services provided in institutional settings, such as nursing homes, are not included in the KUHR system.

Mental health outcomes are measured through registered visits to health care services in the KUHR database using the aforementioned codes. We created two main binary variables indicating whether an individual had at least one visit to primary care services in a given year registered with either a psychological symptom or diagnosed mental disorder. Additional binary variables are created for specific disorders and symptoms with a general prevalence of at least one percent, such as depression, anxiety, and sleep problems.

#### Educational attainment

Data on educational attainment was obtained from Norway’s National Education Database (NUDB), operated by Statistics Norway. The Norwegian Standard Classification of Education (NUS2000) is used to group individuals based on their highest level of completed education. We grouped individuals into four categories: lower secondary schooling or less (i.e. compulsory education), upper secondary schooling, bachelor’s degree, and master’s degree or higher.

#### Partnership status

Information on partnership status comes from the Norwegian Central Population Register. The partnership status registered the year an individual turns 45 was used to classify people as either married or non-married at age 45. While the data on partnership includes multiple options (e.g. married, separated, and registered partner), we only consider marriage versus non-marriage.

#### Ethics

This study was approved by the Regional Committees for Medical and Health Research Ethics (REK) in Norway (#2018/434).

#### Statistical Analysis

Prevalence rates for diagnosed mental disorders and psychological symptoms were calculated separately for men and women at each age, stratified by parenthood and partner status at age 45. Rates were also calculated based on age at first birth and number of children at age 45.

To estimate the association between parenthood and mental health, we used logistic regression models with psychological symptoms or mental disorders as the primary outcome variables. In our full model, we also include covariates that could confound the relationship between fertility and mental health. These include age, educational attainment, partnership status at age 45, and the year of observation. As both fertility behaviour and prevalence of mental illness vary between gender, we analysed men and women separately. Consequently, mental health outcomes are modelled as a function of parenthood though the following two logistic regression equations:

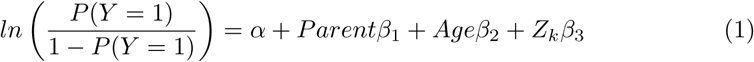

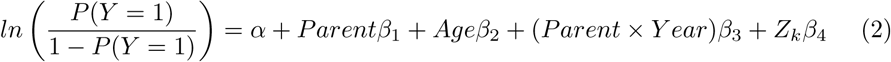

In both equation 1 and 2, *Y* is either a mental disorder or symptoms, and *P* (*Y* = 1) is the probability that a mental disorder or symptoms are present. The full expression on the left-hand side is consequently the log-odds of the probability of Y being equal to 1. *α* is the estimated intercept, *Parent* is a binary variable equal to 1 if one has children at 45, and *Z_k_* is a vector of the remaining control variables (i.e., marital status, educational attainment, year, age at first birth). Our main coefficient of interest is *β*_1_. In equation 2, we have also incorporated an interaction term between *Parent* and *Y ear*. These logistic regression models are estimated for our population analysis. In order to further control for genetic and shared environmental factors between siblings, the same model is also estimated through conditional logistic regressions for our sibling-matched and monozygotic twin sub-samples.

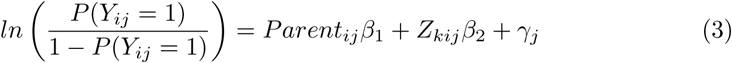

Here, *P* (*Y_ij_*= 1) is the probability that the dependent variable *Y* is equal to 1 for individual *i* in sibship *j*. The full expression in the left-hand side is the log-odds of the probability of *Y_ij_* equalling 1. *Parent_ij_* is again a binary variable for individual *i* in sibship *j*, that equals 1 if one is a parent. *γ_j_* represents the stratum-specific effects, i.e., the individual intercepts for each sibling or twin group.

All statistical analyses were done using Stata MP17 with the logistic and clogit commands.

## 3 Results

The proportion of childless individuals at age 45 was higher among men (16.2%) than women (9.9%). Descriptive statistics for the entire population as well as a yearly sub-sample of 45-year-olds are presented in Table A1.

### Mental health and fertility across adulthood (RQ1)

Figure 1 shows the unadjusted relationship between parenthood at age 45 and mental health over the adult life course for men and women. Parents had a lower risk of mental disorders across all ages. The difference appeared greatest earlier in adulthood and then reduced at older ages. The difference in psychological symptoms was smaller between parents and non-parents. Fathers had a lower prevalence of symptoms throughout the majority of adulthood, whereas mothers had higher levels of symptoms than the childless after age 40.

**Fig. 1.**
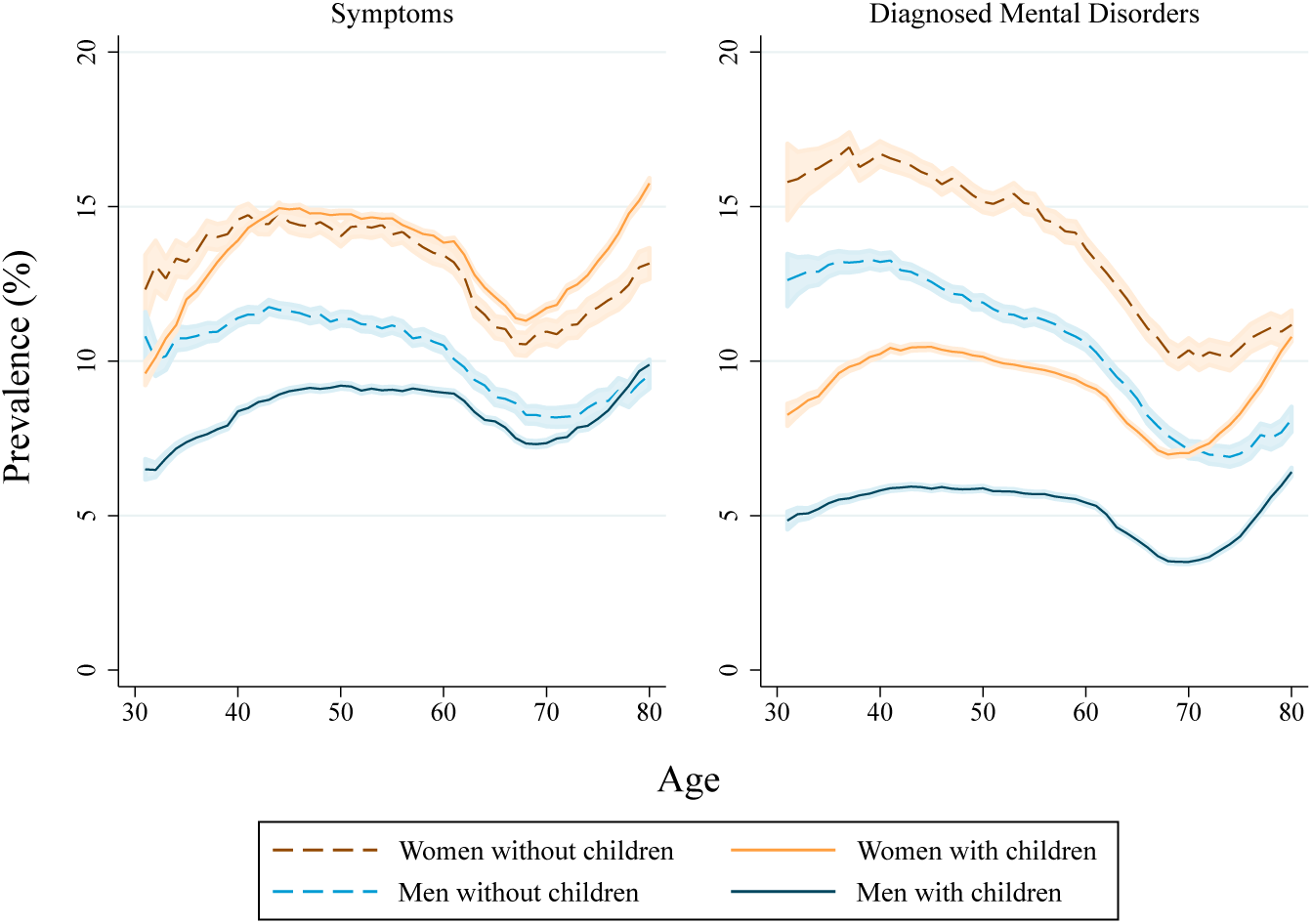
Prevalence of mental disorders and mental symptoms by age, gender, and parenthood at age 45. Shaded areas represent 95% confidence intervals.

Figure 2 presents the estimated associations between parenthood and mental disorders, as well as parenthood and symptoms of disorders/complaints related to mental health. Detailed results are presented in the supplementary materials (Table A2 and A3). We found a significant relationship between mental disorders and parenthood in all of the eight specified models. In both the bivariate and adjusted population models, we found that parents had lower odds of mental disorders. In the fully adjusted model, mothers had 33% lower odds of disorders, whereas fathers had 40% lower odds of disorders. In the sibling-matched analysis we found that mothers had on average 41% lower odds of mental disorders (95% CI 0.58-0.60), while fathers had 44% lower odds (95 % CI 0.55-0.57). When limiting our sample to only monozygotic twin pairs who were discordant in terms of mental disorders, we still found significantly lower odds of mental disorders for both fathers (OR 0.71) and mothers (OR 0.62).

**Fig. 2.**
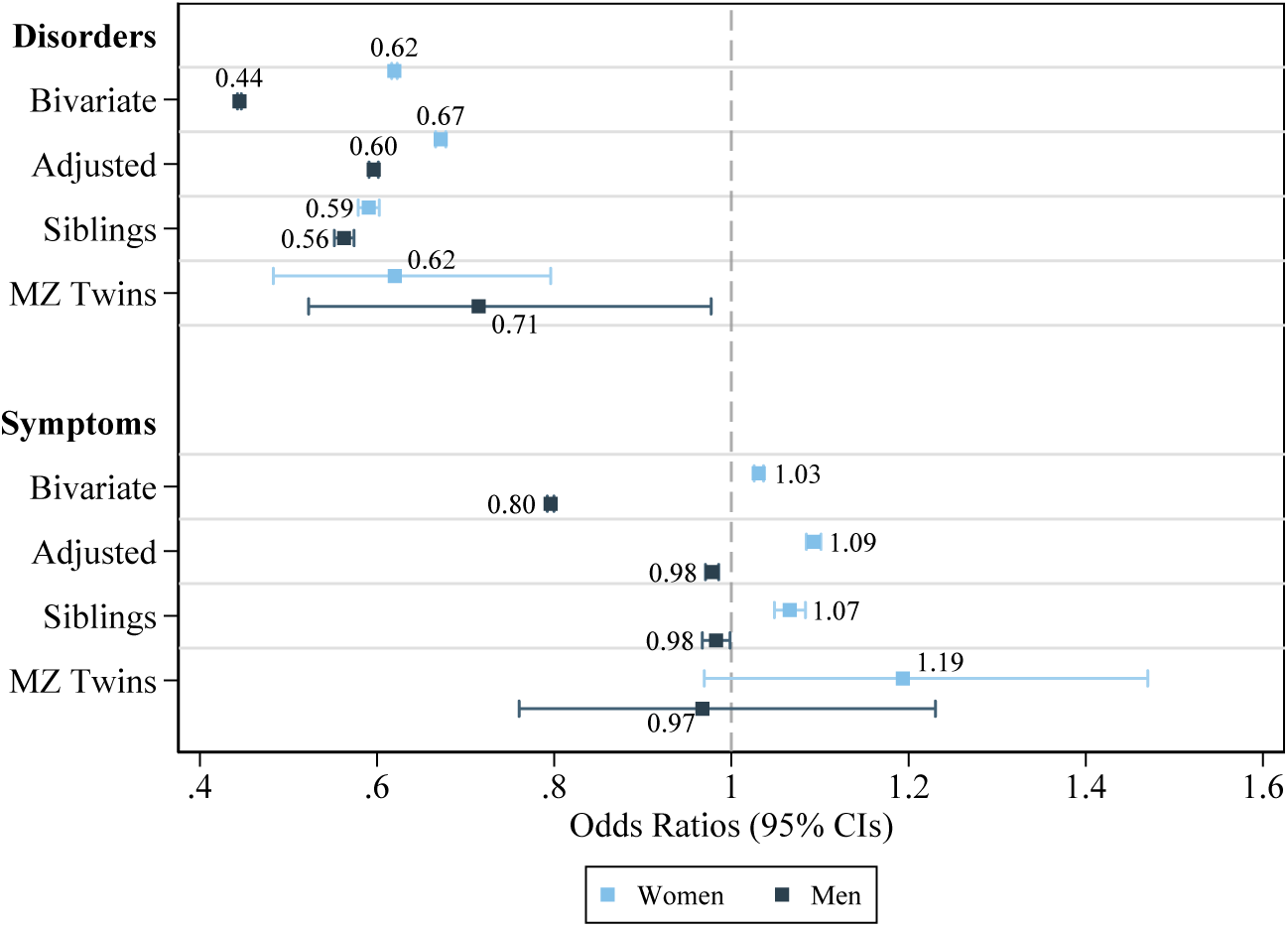
Summary of the different association models estimated between parenthood (parent=1) and mental health.

We also found a significant relationship between mental symptoms/complaints and parenthood in all but two of the eight specified models. However, these went in the opposite direction for men and women. In the fully adjusted population model we found that mothers had 9% higher odds of symptoms, while fathers had 2% lower odds of symptoms compared to the childless. In the sibling-matched analyses we found that mothers had 7% higher odds of symptoms (CI 1.05-1.08), while fathers had 2% lower odds (CI 0-97-0.99). When limiting our sample to only discordant monozygotic twin pairs, we found no significant difference in the odds of symptoms. Nonetheless, the direction of the associations remained consistent with the other models, though with reduced precision in the estimates.

When estimating the same fully adjusted models as in Fig. 2 stratified by age, we found similar results (see Fig. A1 in supplements for detailed results). After adjusting for factors such as education or partnership, parents had significantly lower risk of mental disorders across all ages. For both men and women, the largest difference in terms of mental disorders was seen early in adulthood. Furthermore, we found that especially mothers were prone to psychological symptoms, with an increased risk of symptoms present after age 40. Unlike in the the unadjusted analyses, fathers also had an elevated likelihood of psychological symptoms, which appeared at later ages (ages 55 and above).

### Variations in associations by marital status, socioeconomic status, number of children, and age at first birth (RQ2)

As shown in Figure 3, panel A, the prevalence of symptoms and mental disorders was higher among unmarried individuals, and the difference between parents and non-parents was reduced when taking this into account. In panel B, we see how childless men and women with only compulsory education stand out, with a much higher prevalence of mental disorders compared to the other groups. However, at older ages, this raw difference in mental disorders diminishes among both men and women. In panel C, we see that individuals with two or more children have practically the same risk of mental disorders, whereas individuals who only have one child by age 45 have a higher prevalence of mental disorders throughout life, although not as high as childless individuals. Lastly, panel D shows how the prevalence of mental disorders varies depending on the timing of one’s first birth. Among women, the prevalence of mental disorders is more similar when comparing childless women and mothers who had their first child before the age of 25. On the other hand, among men, fathers who became parents at an early age (before age 27) are more like the other parent-groups than the childless.

**Fig. 3.**
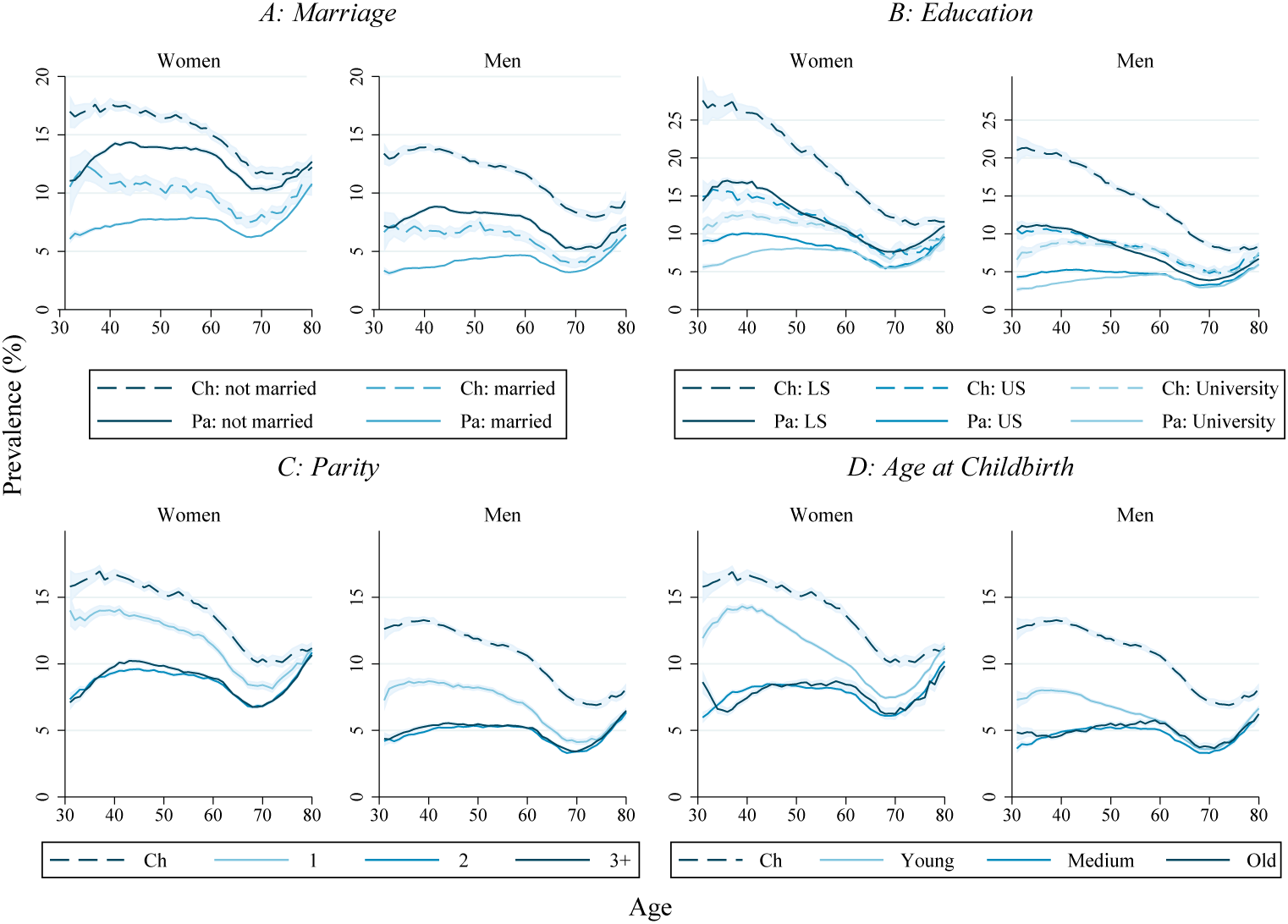
Prevalence of mental disorders (in percent) across age by parenthood and other socioeconomic factors. Ch: childless, Pa: parent. Shaded areas represent 95% confidence intervals. A: Marital status (married vs. non-married) B: Educational attainment (LS: compulsory schooling, US: upper secondary, university) C: Number of children D: Age at first birth

Next, we studied associations between parenthood at age 45 and mental health in different educational groups. The results are shown in Figure 4. Parenthood was associated with a lower risk of mental disorders in all educational groups, but the difference between parents and non-parents was largest among those with only compulsory education (OR 0.62 for women and 0.56 for men). For symptoms, the associations were smaller and did not uniquely run in one direction.

**Fig. 4.**
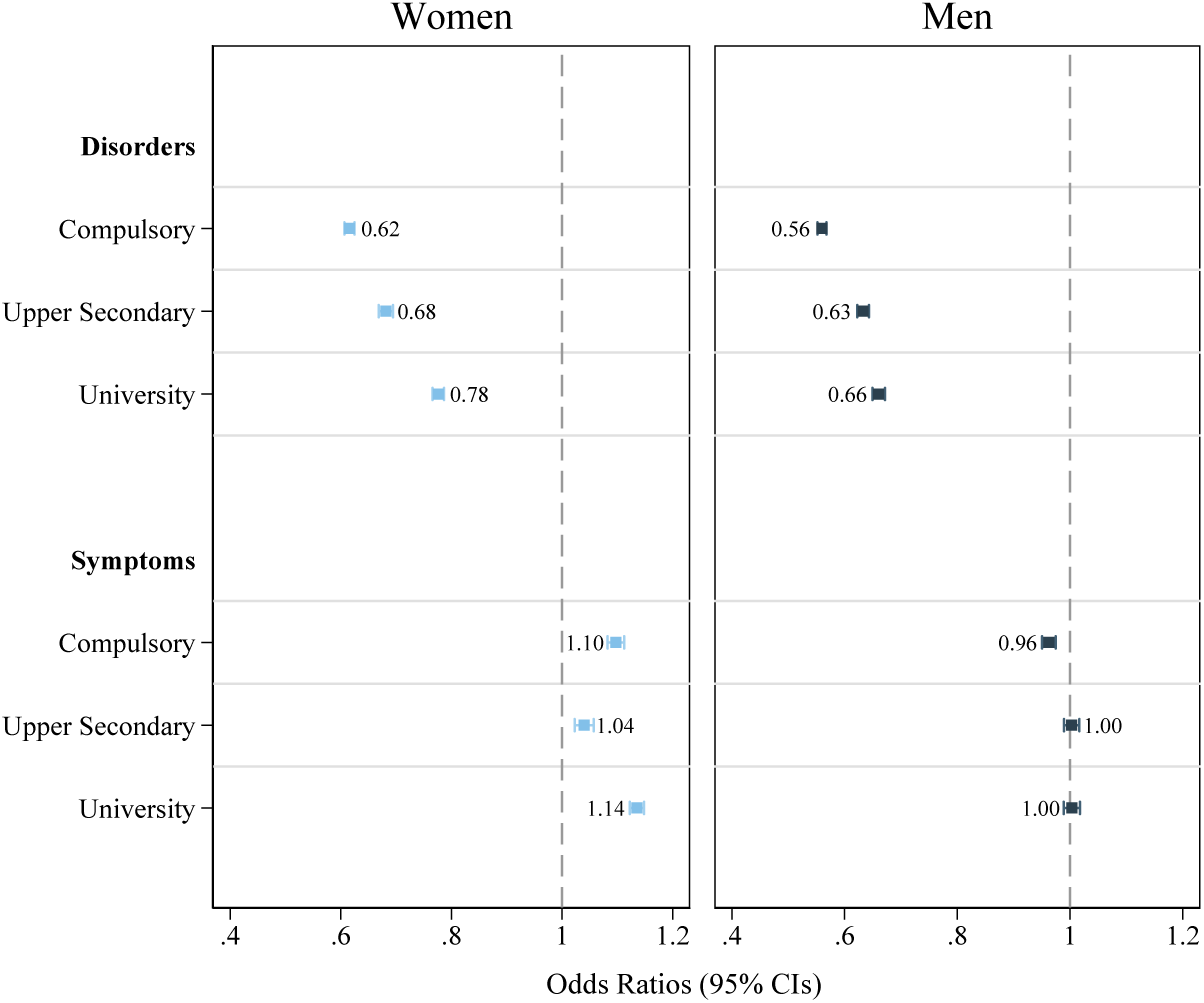
Estimated odds of mental disorders or symptoms depending on parenthood (parent=1) by age 45 for different educational levels. Fully adjusted models, controlling for age, year, age at first birth, and marital status.

We found similar results when looking at the other three markers linked to socioeconomic status (marriage, number of children, and age at childbirth). The odds of having a mental disorder was always higher among childless individuals. The odds of mental disorders was lowest for parents relative to the childless when looking at unmarried individuals or when focusing on parents with more than one child. Parents who had their first child at an early age were more similar to the childless, however, there still remained a significant difference in the risk of mental disorders. With regards to symptoms, the associations were less uniform. Detailed results are presented in Figure A2.

### Changes over time (RQ3)

As shown in Fig. 5, when looking at mental health over time, we see how the association between mental health and parenthood persists across our study period. It is also evident that while the prevalence of mental disorders remained relatively stable for parents of two or more children, there was a steady increase in the prevalence of mental disorders among both the childless and parents of only children. This increase in mental disorders was especially prominent for childless males. Incorporating interaction terms between parenthood and year in our models (see eq. 2) revealed a statistically significant change in how parenthood and the likelihood of mental disorders were related. Fig. 6 shows how the gap in likelihood of a mental disorder between parents and the childless has grown from 2006 to 2019, even after adjusting for other factors such as educational attainment or marital status. The gap in mental health widened more for men than women. In 2006, fathers were on average around 5 percentage points less likely to have a mental disorder compared to childless men. However, in 2019, this difference had grown to around 7 percentage points. We also studied how these changes over time differed across levels of educational attainment. While the mental health gap grew in all groups, it grew the most among men and women with only compulsory education (see Fig. A3 and A4 in supplements for details).

**Fig. 5.**
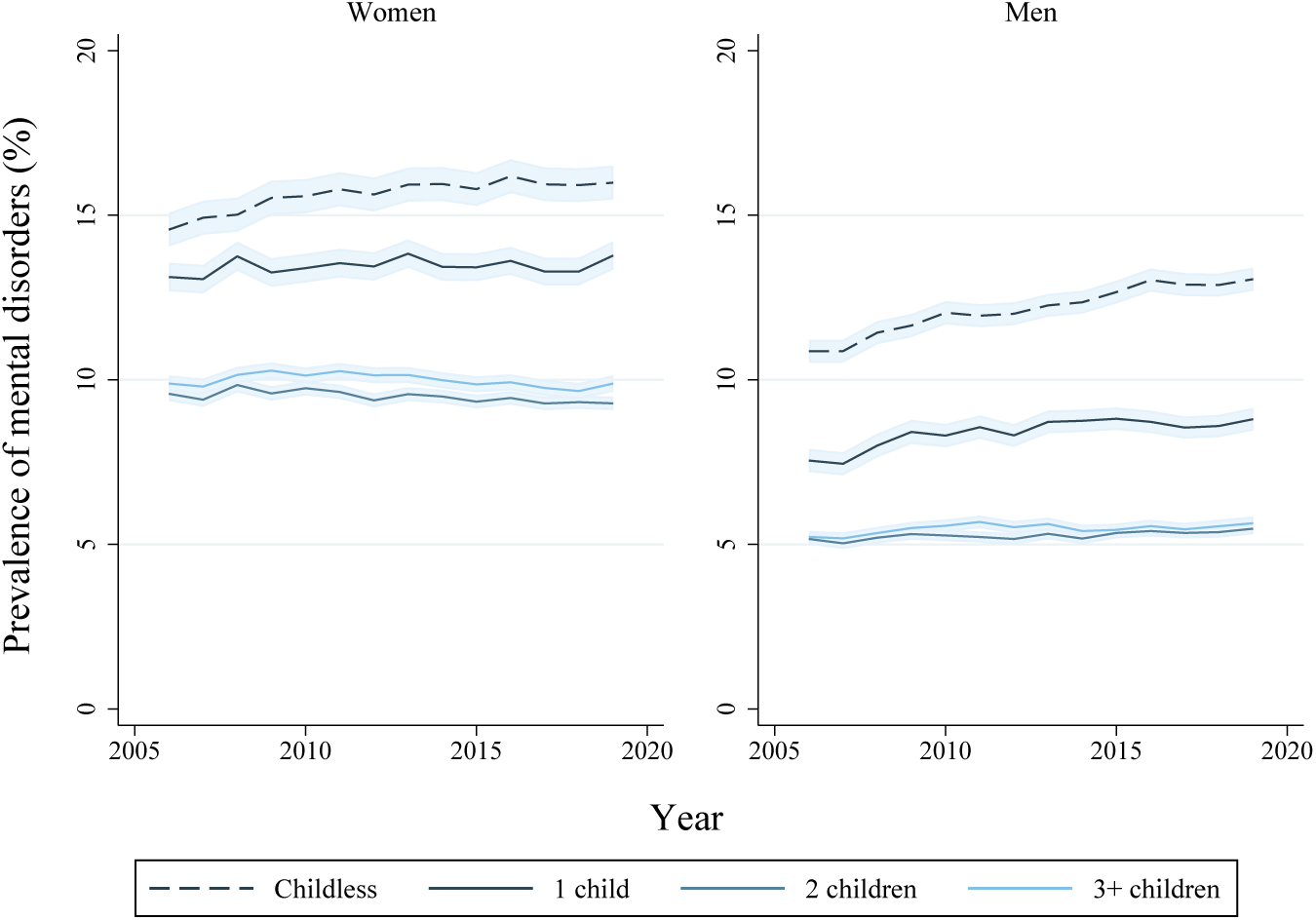
Prevalence of mental disorders among men and women aged 45-50 by parenthood, from 2006 to 2019. Shaded areas represent 95% confidence intervals.

**Fig. 6.**
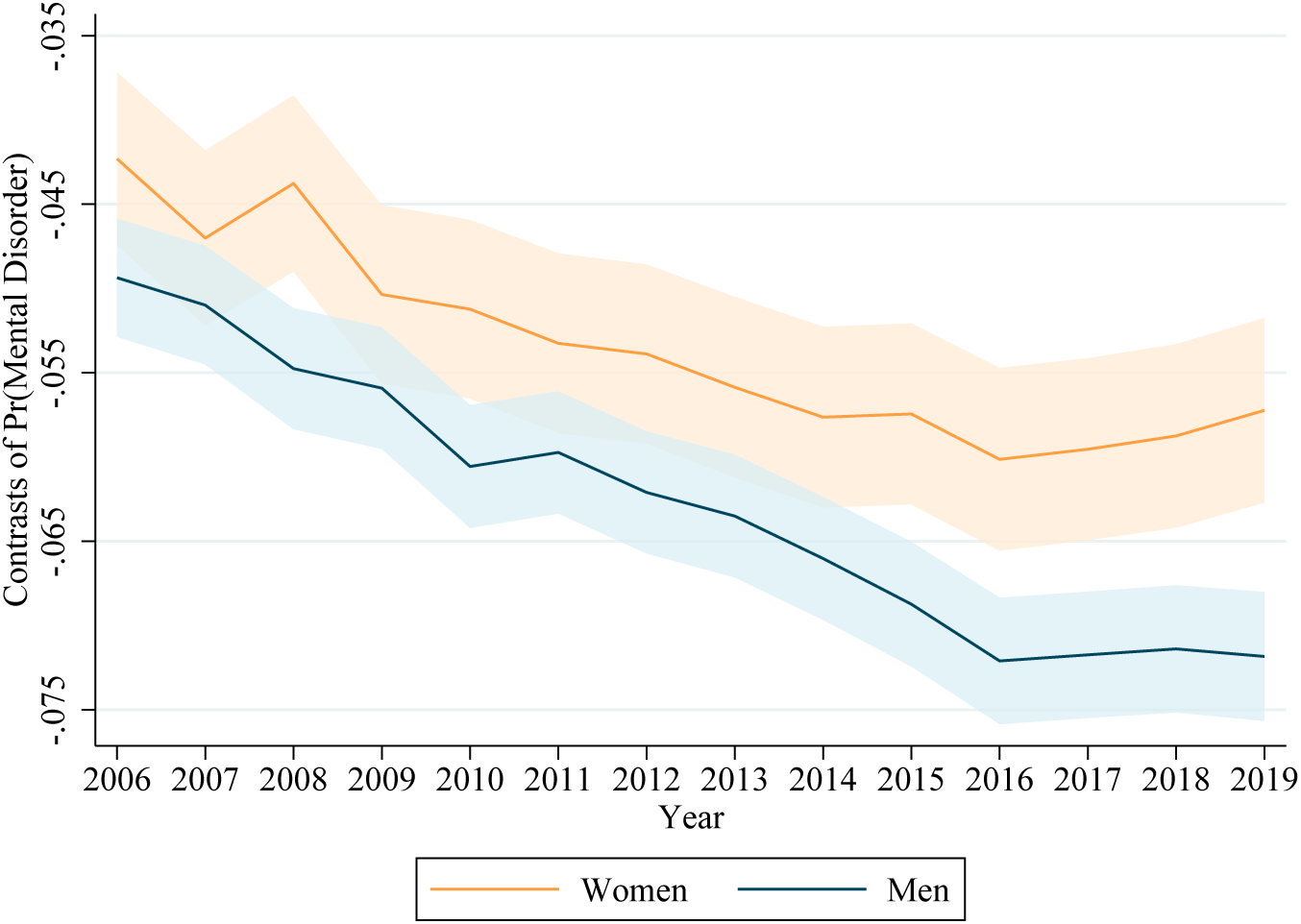
Contrasts of predicted probabilities of a mental disorder (parent=1) with 95% confidence intervals. Models are fully adjusted and control for age, marital status, age at childbirth, and educational attainment. The dataset is limited to ages 45-50 (inclusive) to avoid results driven by compositional changes.

### Specific diagnoses and symptoms (RQ4)

Figure 7 shows the prevalence of specific diseases and symptoms among parents and non-parents. We have focused on diseases or symptoms with a significant presence (*>* 1%) in most age groups. We see that specific ICPC-2 codes are differently associated with parenthood. For example, while the childless had on average a higher prevalence of depressive and anxiety disorders, there was a less marked difference when looking at feelings of depression. Conversely, the childless still had a higher prevalence of feelings of anxiety, especially among men. On the other hand, parents, and especially mothers, had a higher prevalence of acute stress reactions compared to childless adults. We then estimated the association between each symptom or diagnosis and parenthood separately, with the results presented in Figure 8. In these fully adjusted models, we also found that parenthood was not consistently associated with adverse mental health outcomes. While parents had lower odds of both anxiety (OR 0.78 for women and 0.70 for men) and depressive disorders (OR 0.94 and 0.90), certain symptoms were positively associated with parenthood. Both mothers and fathers had higher odds of feelings of depression (OR 1.19 and 1.09) and acute stress reactions (OR 1.37 and 1.41) when compared to otherwise similar childless adults. For the two other most prevalent symptoms, feelings of anxiety (OR 0.90 and 0.83) and sleep disturbance (OR 0.98 and 0.96), parenthood was again associated with lower odds.

**Fig. 7.**
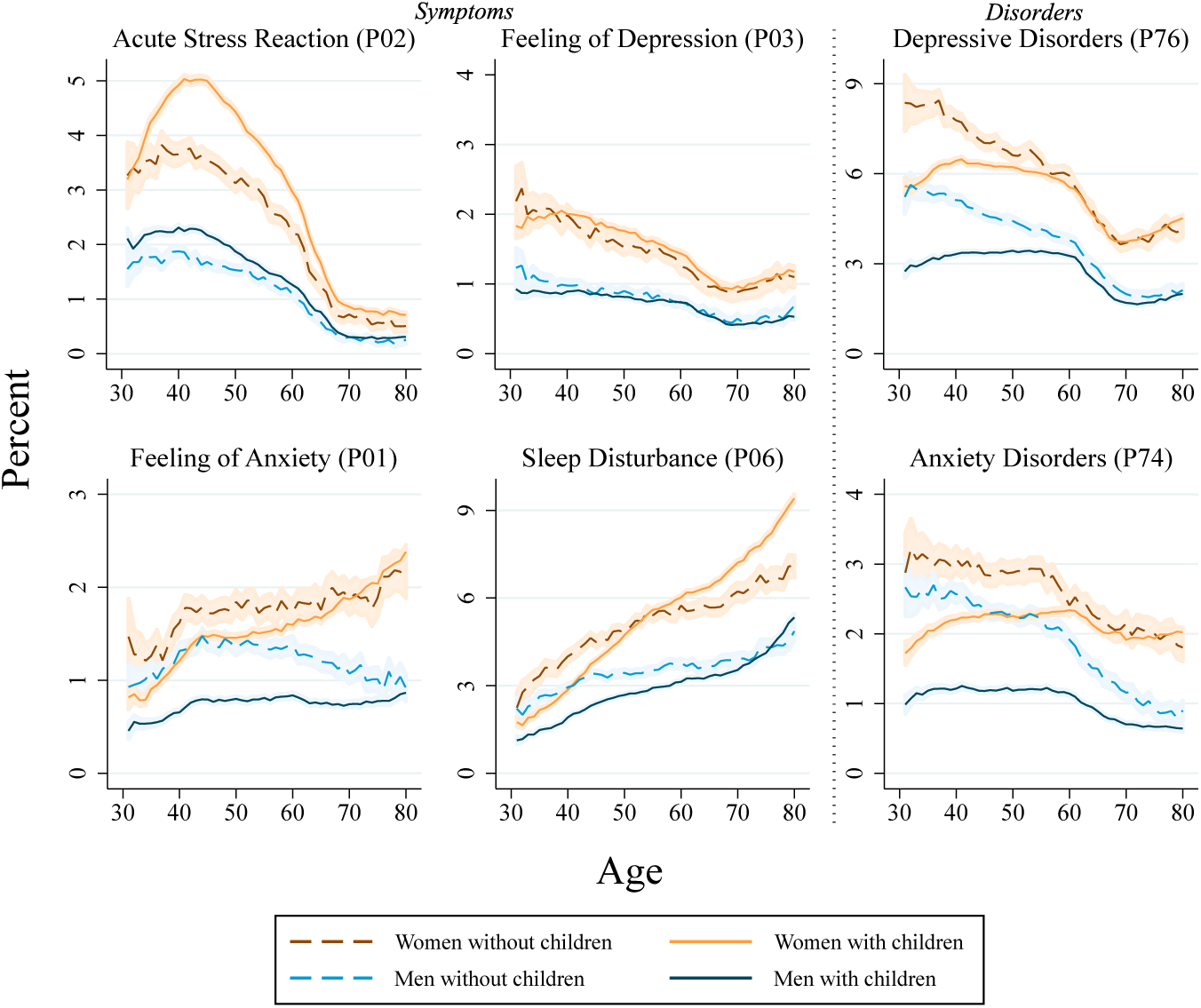
Prevalence of various mental disorders and symptoms by age, gender, and parenthood (at age 45). Shaded areas represent 95% confidence intervals.

**Fig. 8.**
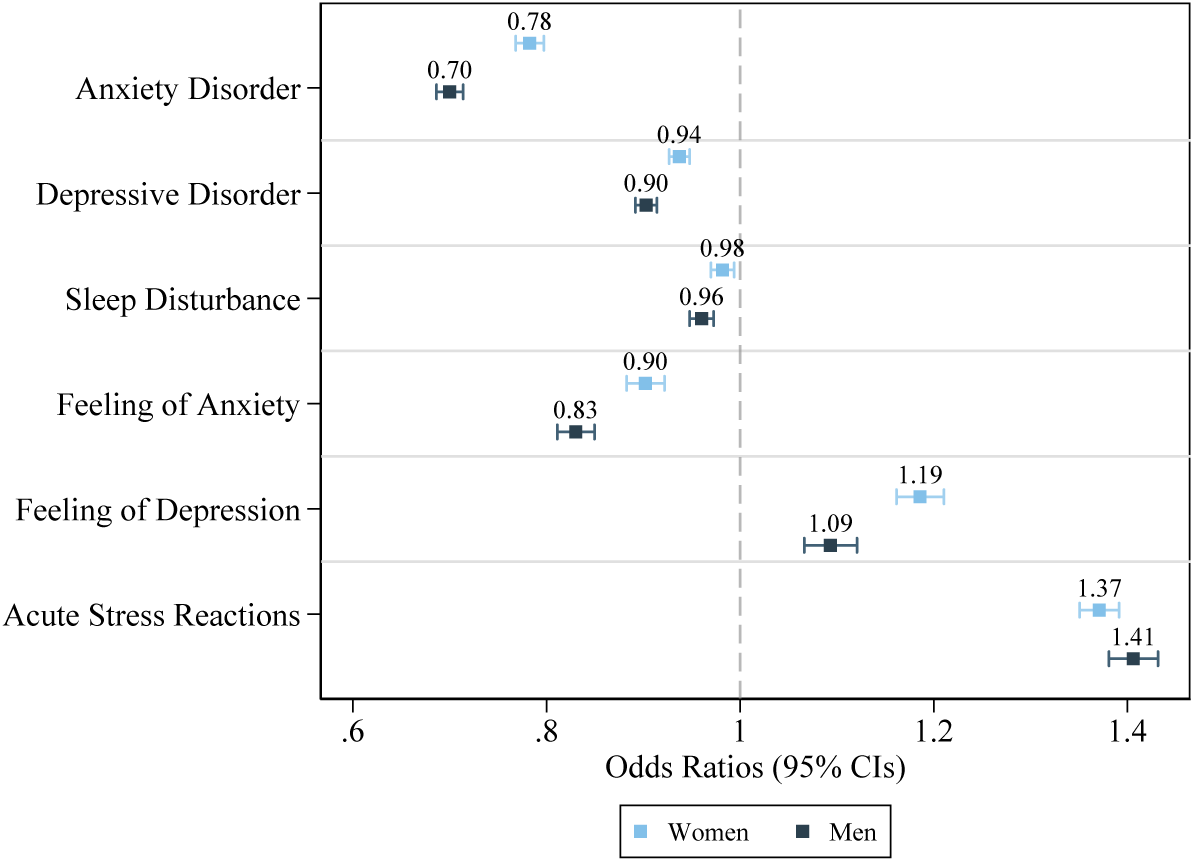
Associations between specific mental disorders/symptoms and parenthood (parent=1). Fully adjusted models, controlling for age, year, age at first birth, education, and marital status. ICPC-2 codes used are P01 (feeling of anxiety), P02 (acute stress reaction), P03 (feeling of depression), P06 (sleep disturbance), P74 (anxiety disorder), and P76 (depressive disorder.)

## 4 Discussion

We have studied the association between parenthood and mental health outcomes for the entire Norwegian-born population aged 31-80 years old between 2006 and 2019. In addition to our main population analyses, we also studied how parenthood and mental health outcomes were associated within same-sex sibling sets and monozygotic twin pairs. We found that parents had a lower likelihood of mental disorders. At the same time, mothers had a slightly higher risk of symptoms/complaints linked to mental health, while fathers had an almost negligible reduction in risk. The difference in mental health between parents and non-parents was present throughout adulthood but diminished at older ages. The parenthood-gap in mental health was evident in all sub-groups of the population, as defined by educational attainment, partnership status, and timing of first birth. We also found that the difference in mental health between the childless and parents increased during our study period. The difference in mental health grew more among men and those with the lowest level of educational attainment. With regards to specific diagnoses and symptoms, we found that parenthood was associated with a lower risk of depressive disorder, anxiety disorder, sleep disturbance, and feeling of anxiety. On the other hand, parenthood was associated with a higher risk of feeling of depression and acute stress reactions.

Previous research on the link between parenthood and mental health has found mixed results [e.g. 12, 13, 15, 28–30]. We found that both male and female parents had considerably lower risk of mental disorders compared to childless adults. The difference in odds was especially prominent early in adulthood, but parents remained at lower risk even at advanced ages. There is little pre-existing research examining how the association between parenthood and mental disorders may change across age. Our study indicates a clear mental health advantage for parents throughout life. This association also remains after adjustment for observed confounders like education or partnership. Hence, it cannot easily be dismissed as purely driven by socioeconomic factors. Moreover, the association remained significant among discordant monozygotic twins, indicating that a relationship exists independently of the familial part of unobserved confounders. Having children may improve life quality, or mentally healthy individuals may be more prone to selection into parenthood [7]. Regardless of the direction of effects, the gap in mental health between childless individuals and parents suggest that childless adults need more health care services throughout life. With an ageing population and an overall increased use of health services, the additional health care expenditures associated with childlessness may not be negligible.

We also found that the gap in mental disorders narrowed throughout adulthood into older ages. One possible cause of this diminishing gap is the competing risk of death and consequently some survivorship bias [31]. It is possible that childless individuals who live longer may be overall healthier than the childless who die early, and we know that mortality rates are overall higher for the childless [18]. Second, there might be differences in help-seeking behaviour that develop differently over time for the parent and non-parent groups. The presence of children or other family-members may result in increased help-seeking among elderly parents [32], which could increase the reported cases of mental disorders or symptoms. Furthermore, some parents may also experience disability, illness or death among their children, which negatively impacts their mental health [33, 34]. Lastly, as children become adults and leave the household, it is possible that parents rely more on other arenas for their social network. Consequently, the difference in mental health between parents and the childless may be smaller when only empty-nesters (i.e. parents at older ages) are analysed.

We also found significant differences between parents and the childless when studying specific social groups. While education and partnership played an important role in attenuating the relationship between parenthood and mental health outcomes, we found a significant association between parenthood and mental disorders across all groups of educational attainment. However, for both men and women we found the largest difference in odds of mental disorders among those with the lowest level of educational attainment. One possible explanation for this difference could be that there are differences in the degree of involuntary childlessness across the various groups. Men, and especially women with higher education have previously been seen as more prone to actively opt out of having children due to prioritising their career [25]. However, more recent studies have found that the educational gradient in childlessness has flipped among Norwegian women belonging to more recent birth cohorts [26]. Consequently, the strong association between parenthood and mental health found among those with low levels of education may be driven by other factors, such as a greater importance of the change in social networks and capital parenthood may bring [35, 36]. Additionally, we have studied how the mental health of parents and childless adults have evolved across our study period. While the prevalence of mental disorders remained relatively stable for men and women with two or more children, we found an especially large increase in mental disorders among childless men. Incorporating interaction variables between parenthood and year in our analysis revealed significant interaction effects. Estimating predicted prevalences of mental disorders across our study period also showed an increased gap in mental disorders between parents and the childless. To our knowledge, there are currently no larger studies investigating how the association between mental health and parenthood has evolved over time. However, our results suggest that parenthood has become an increasingly important marker of mental health in the Norwegian population. There are multiple possibilities for why this could be. One reason is that childlessness is becoming more involuntary, either due to lower fecundity or an increased difficulty in finding a partner. This stronger selection into parenthood may then increase the observed difference in mental health between parents and non-parents. Given that we find less change in the mental health of parents, but a marked worsening of the mental health of the childless, this explanation seems likely.

Lastly, we studied how the mental health of parents and non-parents differed depending on the symptoms or diagnoses studied. Parenthood was not associated with a lower occurrence of all negative mental health outcomes. While parents on average had a lower likelihood of mental disorders, mothers had a slightly higher chance of having any symptoms related to mental illness. Fathers had a slightly lower risk of symptoms. However, this difference in symptom occurrence between fathers and childless men was very small. Given that women still tend to face a larger share of the household and child-rearing responsibilities [37], one could hypothesise that this uneven burden results in higher occurrence of psychological symptoms among mothers.

When focusing on specific symptoms and diagnoses, we also showed that parenthood was not always associated with better mental health. Certain symptoms, like feelings of depression and acute stress reactions were more common among men and women who were parents. Our diverging findings with regards to these specific symptoms could explain why the overall findings on parenthood and symptoms were small in magnitude. These diverging associations also illustrate why it is important to separate symptoms from the more severe disorders, as well as look at specific disorders and symptoms. Combining multiple outcomes into one simple measure of mental health could hide the true variability in associations. This may also contribute to explaining the diverging results in previous studies that have used different operationalisations of mental health [12, 13, 15, 28–30].

In our study, we had no way to observe an individual’s fertility intentions, and consequently, we could not separate involuntary and voluntary childlessness. We therefore believe that further research into how mental health may be differently associated with childlessness depending on fertility intentions would be beneficial to knowledge on this topic.

Our study has several strengths. First and foremost, we have complete coverage of primary health care data and accurate fertility data for an entire population. Second, the detailed health data means we can look at specific diagnoses and symptoms, which allows us to identify the diseases or symptoms most strongly associated with parenthood. Finally, due to the richness of the register data, we can also use both a sibling- and twin-matched design to limit the confounding influence of genetics and shared family environments.

While the use of complete registry data in this analysis means that our findings relate to the majority of the Norwegian population, this study still has a few limitations. First, while the primary care database reflects the health of most of the Norwegian population, health care services provided at nursing homes and institutions are not registered. This means that especially for the oldest age groups, we will underestimate prevalence rates. Furthermore, there might be differences in how a practitioner evaluates a patient’s symptoms depending on factors such as socioeconomic status or gender, which could impact our results in ways we cannot control for. Due to the deterministic relationship between age, year, and birth cohort we also cannot fully disentangle period, cohort, and year effects in this study. Lastly, as we have limited our study to Norwegian-borns, we cannot say anything about how parenthood and mental health may be related among Norwegians born elsewhere.

We have studied the association between parenthood and mental health in the Norwegian population. We found that there are significant associations between parenthood and mental health outcomes. Even after adjusting for important confounders, parents had significantly better mental health than the childless. Sex-stratified analyses also revealed a larger disparity in mental health between fathers and childless men compared to women. And importantly, the gap in mental health between the childless and parents has increased during our study period. A growing gap signals that either selection into parenthood has become stronger over time, or that remaining childless has become worse for one’s mental health, or both. Thus, the increased association between parenthood and mental health could be due to both stronger partner-selection among those wanting children and more involuntary childlessness. Our findings have important implications. If childlessness increases the risk of mental disorders, its increasing occurrence could imply a decline in the mental health of the population. Furthermore, if poor mental health reduces the likelihood of becoming a parent, policy aimed at encouraging parenthood should not ignore the importance of mental health in childhood and adolescence. Finally, policy makers should be aware of the mental health gap between parents and non-parents.

## Data Availability

The data used in this study include primary health care records, demographic information, and educational outcomes for entire cohorts of the Norwegian population. Therefore, the data used in this study is only available by application to the Regional Committees for Medical and Health Research Ethics and the data owners (the Norwegian Directorate of Health and Statistics Norway). The authors cannot share these data with other researchers due to its sensitive nature and potential for identification. However, researchers can contact the authors if they have questions about the data or have overlapping research projects.

## Acknowledgements

This work is part of the PARMENT project and was funded by the Research Council of Norway (#334093). This work was partly supported by the Research Council of Norway through its Centres of Excellence funding scheme, project number 262700 and co-funded by the European Union (ERC, BIOSFER, 101071773). Views and opinions expressed are however those of the author(s) only and do not necessarily reflect those of the European Union or the European Research Council. Neither the European Union nor the granting authority can be held responsible for them.

## Appendix A Supplements

**Fig. A1.**
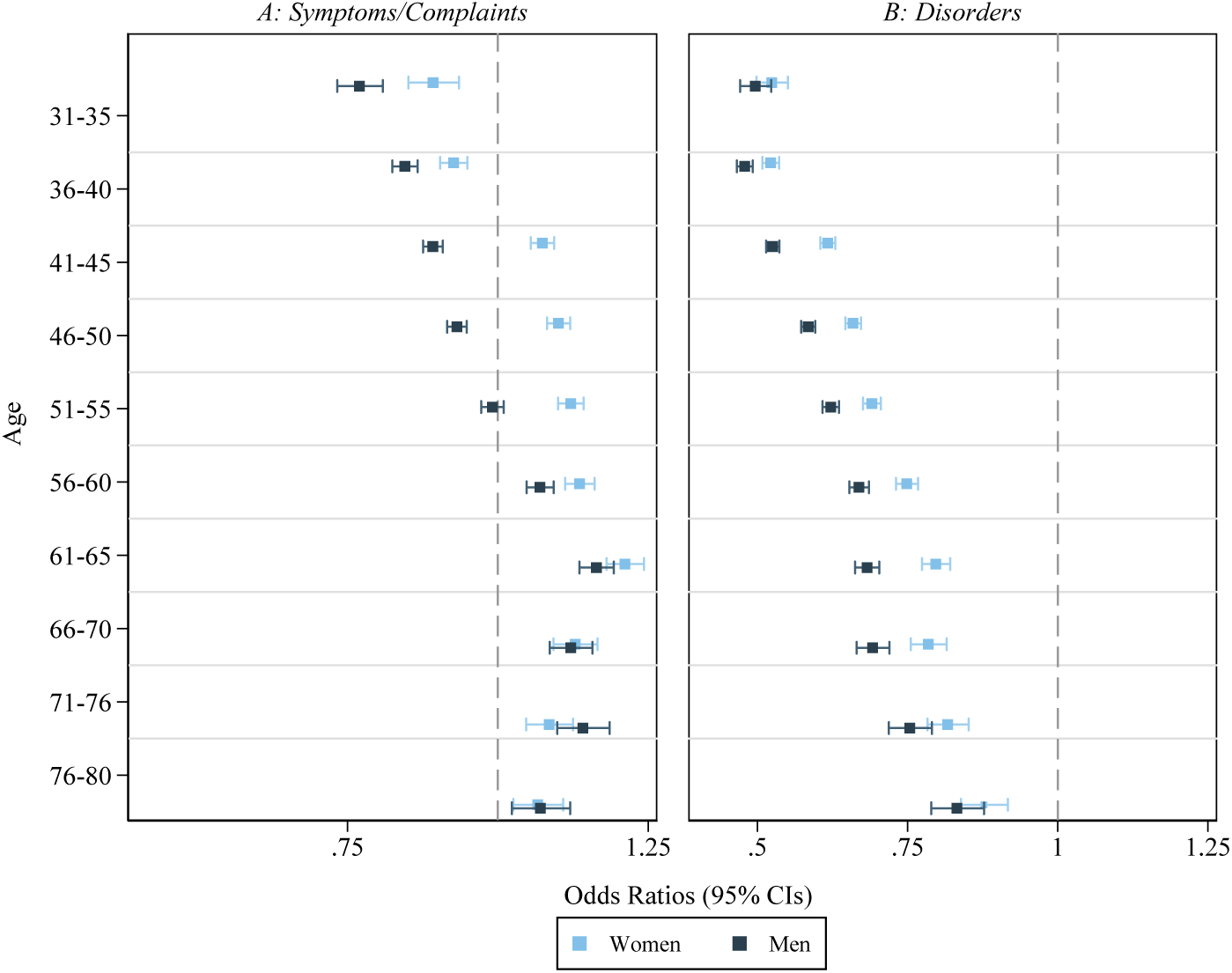
Estimated odds ratios of psychological symptoms/complaints (A) and mental disorders (B) by parenthood (=1) at age 45, controlling for educational attainment, partnership, age at first birth, and year.

**Fig. A2.**
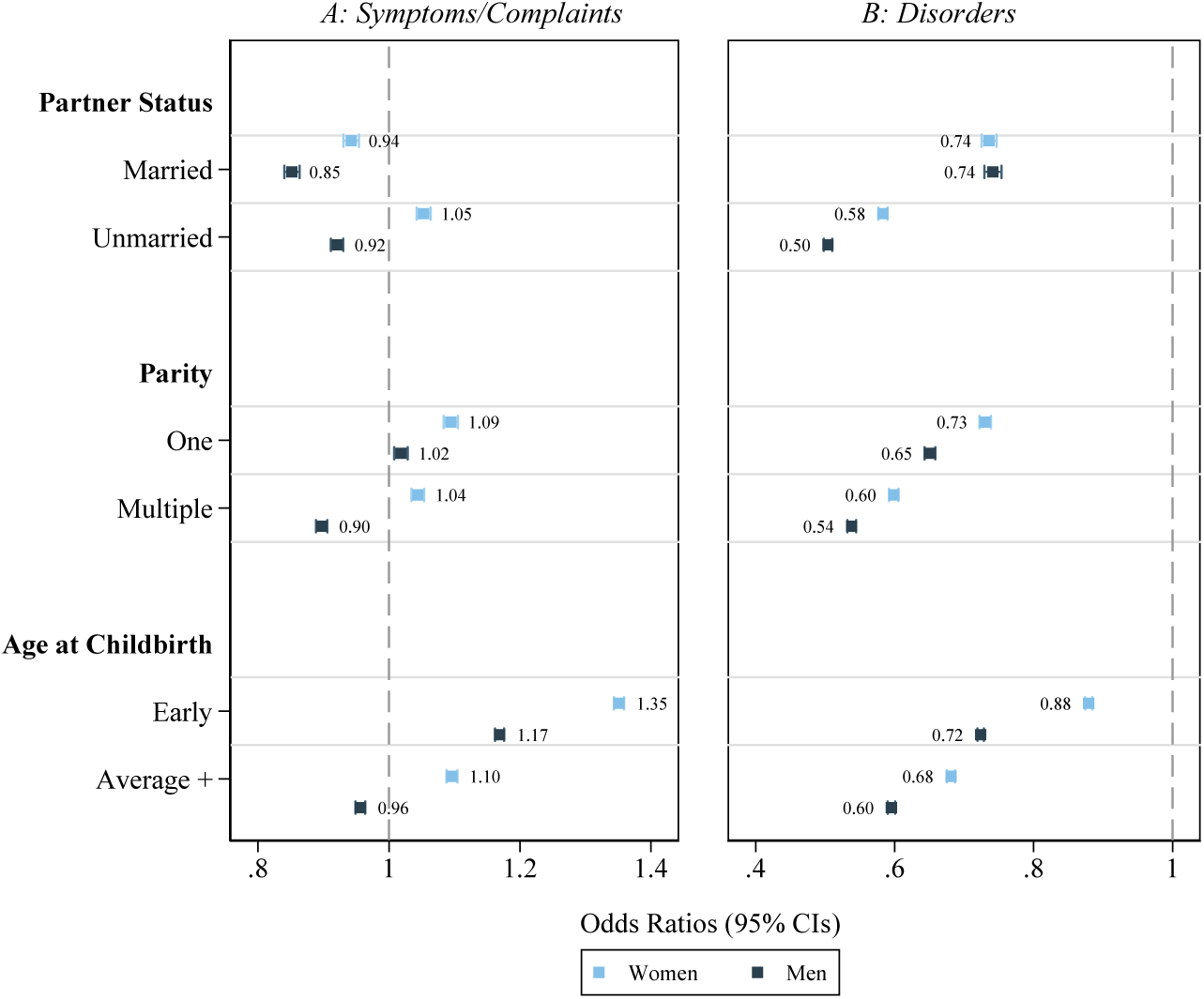
Estimated odds ratios of psychological symptoms/complaints (A) and mental disorders (B) by parenthood (=1) at age 45, controlling for educational attainment, age, and year. Models also control for the other markers of socioeconomic background not used to limit the sample (i.e. partner status, parity, and age at childbirth).

**Fig. A3.**
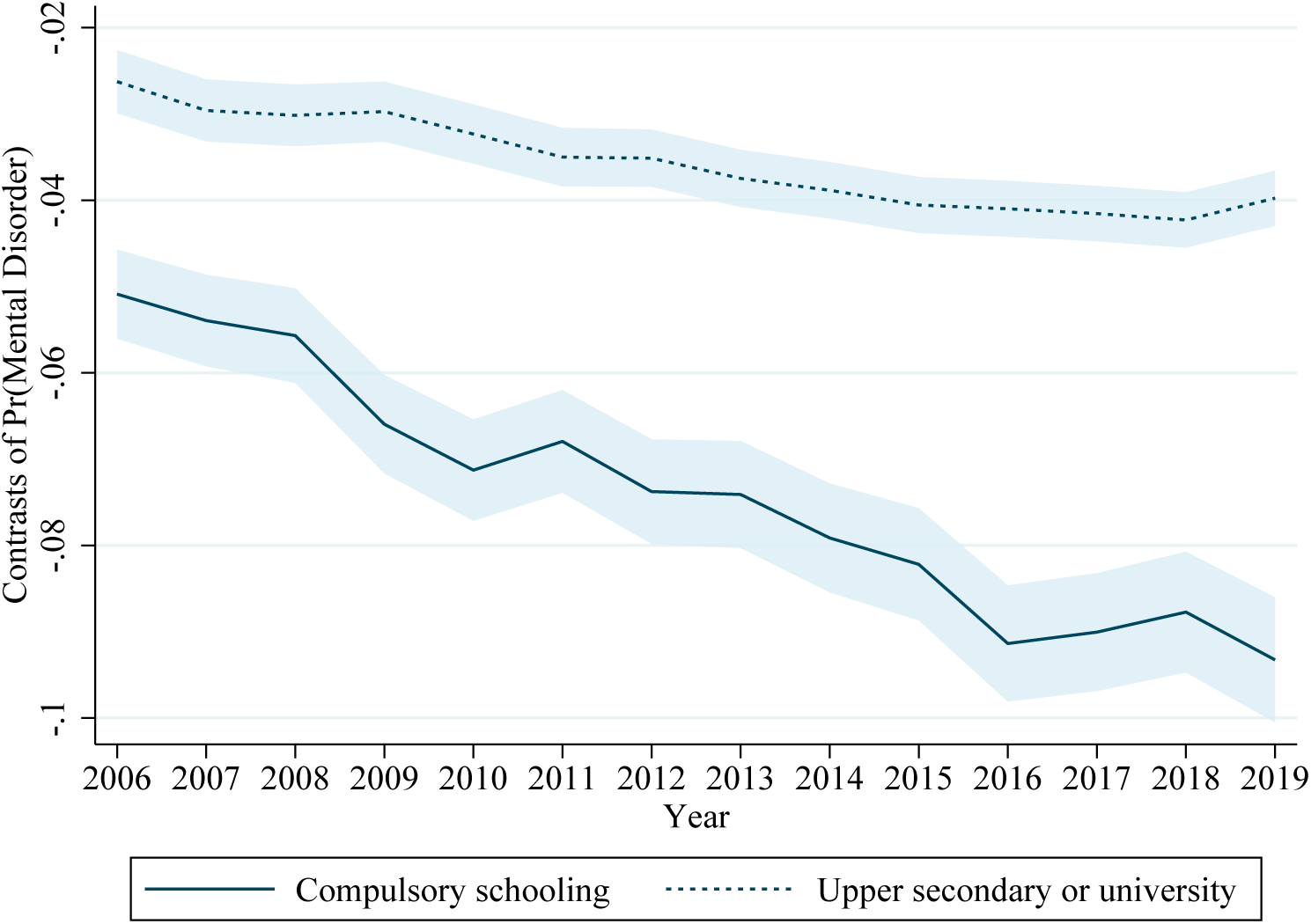
Contrasts of predicted probabilities of a mental disorder (parent=1) with 95% confidence intervals. Models are fully adjusted and control for gender, age, marital status, and age at childbirth. The dataset is limited to ages 45-50 (inclusive) to avoid results driven by compositional changes.

**Fig. A4.**
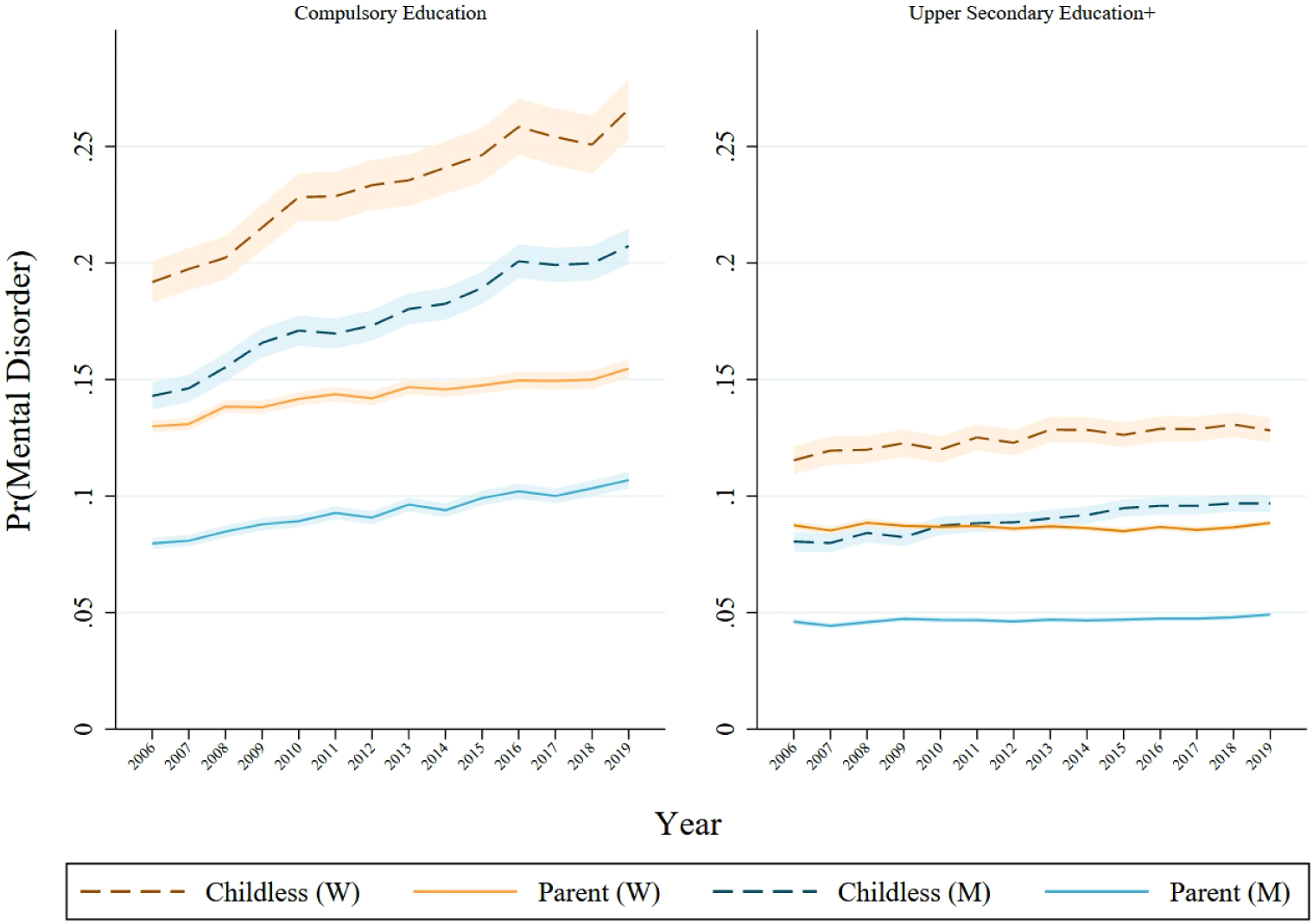
Predicted probabilities of a mental disorder with 95% confidence intervals. Models are fully adjusted and control for age, marital status, and age at childbirth. The dataset is limited to ages 45-50 (inclusive) to avoid results driven by compositional changes

**Table A1.** Summary Statistics.

|  | 2006 | 2007 | 2008 | 2009 | 2010 | 2011 | 2012 | 2013 | 2014 | 2015 | 2016 | 2017 | 2018 | 2019 |
| --- | --- | --- | --- | --- | --- | --- | --- | --- | --- | --- | --- | --- | --- | --- |
| **Full sample** |  |  |  |  |  |  |  |  |  |  |  |  |  |  |
| Age | 51.88 | 52.48 | 53.10 | 53.72 | 54.36 | 54.99 | 55.64 | 56.28 | 56.96 | 57.63 | 58.31 | 58.98 | 59.65 | 60.31 |
| Female | 0.50 | 0.50 | 0.50 | 0.50 | 0.50 | 0.50 | 0.50 | 0.50 | 0.50 | 0.50 | 0.50 | 0.50 | 0.50 | 0.50 |
| Childless at 45 | 0.13 | 0.13 | 0.13 | 0.13 | 0.13 | 0.13 | 0.13 | 0.13 | 0.13 | 0.13 | 0.13 | 0.13 | 0.13 | 0.13 |
| Married at 45 | 0.65 | 0.65 | 0.65 | 0.65 | 0.65 | 0.65 | 0.65 | 0.64 | 0.64 | 0.64 | 0.64 | 0.63 | 0.63 | 0.63 |
| Age at First Birth | 26.46 | 26.45 | 26.45 | 26.45 | 26.46 | 26.46 | 26.48 | 26.49 | 26.51 | 26.53 | 26.55 | 26.57 | 26.60 | 26.63 |
| Early Age at First Birth | 0.42 | 0.42 | 0.43 | 0.43 | 0.43 | 0.43 | 0.42 | 0.42 | 0.42 | 0.42 | 0.42 | 0.42 | 0.42 | 0.41 |
| Med. Age at First Birth | 0.36 | 0.36 | 0.36 | 0.36 | 0.36 | 0.36 | 0.36 | 0.36 | 0.36 | 0.36 | 0.36 | 0.36 | 0.36 | 0.36 |
| Adv. Age at First Birth | 0.09 | 0.09 | 0.09 | 0.09 | 0.09 | 0.09 | 0.09 | 0.09 | 0.09 | 0.09 | 0.09 | 0.09 | 0.09 | 0.09 |
| Lower Secondary (Compulsory) | 0.47 | 0.46 | 0.45 | 0.45 | 0.44 | 0.43 | 0.43 | 0.42 | 0.41 | 0.41 | 0.40 | 0.39 | 0.39 | 0.38 |
| Upper Secondary | 0.25 | 0.26 | 0.26 | 0.26 | 0.27 | 0.27 | 0.27 | 0.28 | 0.28 | 0.28 | 0.28 | 0.29 | 0.29 | 0.29 |
| Bachelor's Degree | 0.21 | 0.21 | 0.22 | 0.22 | 0.22 | 0.22 | 0.23 | 0.23 | 0.23 | 0.23 | 0.24 | 0.24 | 0.24 | 0.24 |
| Master's Degree | 0.07 | 0.07 | 0.07 | 0.07 | 0.07 | 0.07 | 0.07 | 0.08 | 0.08 | 0.08 | 0.08 | 0.08 | 0.08 | 0.09 |
| Mental Disorders | 7.86 | 7.86 | 8.18 | 8.22 | 8.24 | 8.17 | 8.07 | 8.18 | 8.1 | 8 | 7.9 | 7.62 | 7.48 | 7.43 |
| Psychological Symptoms | 8.78 | 9.14 | 9.94 | 10.25 | 10.55 | 10.9 | 11.14 | 11.64 | 12.24 | 12.8 | 13.12 | 12.79 | 12.83 | 13.08 |
| Observations | 2,394,217 | 2,352,756 | 2,312,970 | 2,273,388 | 2,234,767 | 2,195,788 | 2,156,854 | 2,117,671 | 2,080,590 | 2,043,811 | 2,006,656 | 1,968,715 | 1,929,826 | 1,889,729 |
| **Aged 45** |  |  |  |  |  |  |  |  |  |  |  |  |  |  |
| Childless (Men) | 18.11 | 18.6 | 18.8 | 18.61 | 19.3 | 18.93 | 18.91 | 19.2 | 18.81 | 19.47 | 19.51 | 19.65 | 20 | 20.28 |
| Childless (Women) | 10.94 | 10.97 | 11.26 | 11.12 | 10.97 | 10.69 | 10.93 | 10.55 | 11.05 | 11.06 | 11.21 | 11.22 | 11.19 | 11.76 |
| Age at First Birth | 27.13 | 27.19 | 27.32 | 27.46 | 27.54 | 27.67 | 27.76 | 27.86 | 27.98 | 28.12 | 28.31 | 28.51 | 28.63 | 28.70 |
| Mental Disorders | 8.62 | 8.53 | 8.97 | 8.90 | 9.06 | 9.35 | 9.00 | 9.20 | 9.23 | 9.04 | 9.24 | 9.08 | 9.22 | 9.66 |
| Psychological Symptoms | 9.33 | 9.55 | 10.50 | 11.05 | 11.07 | 11.97 | 11.67 | 12.18 | 12.93 | 13.43 | 14.00 | 13.76 | 13.92 | 14.63 |
| Observations | 57,092 | 56,916 | 57,898 | 59,995 | 60,466 | 61,400 | 61,238 | 62,396 | 62,870 | 60,042 | 60,939 | 59,905 | 57,194 | 55,694 |

*Summary stats are for the entire sample in that year. For example, age at first birth in 2006 is not calculated only based on those that gave birth in 2006, but for everyone in the sample in 2006.*

**Table A2.**
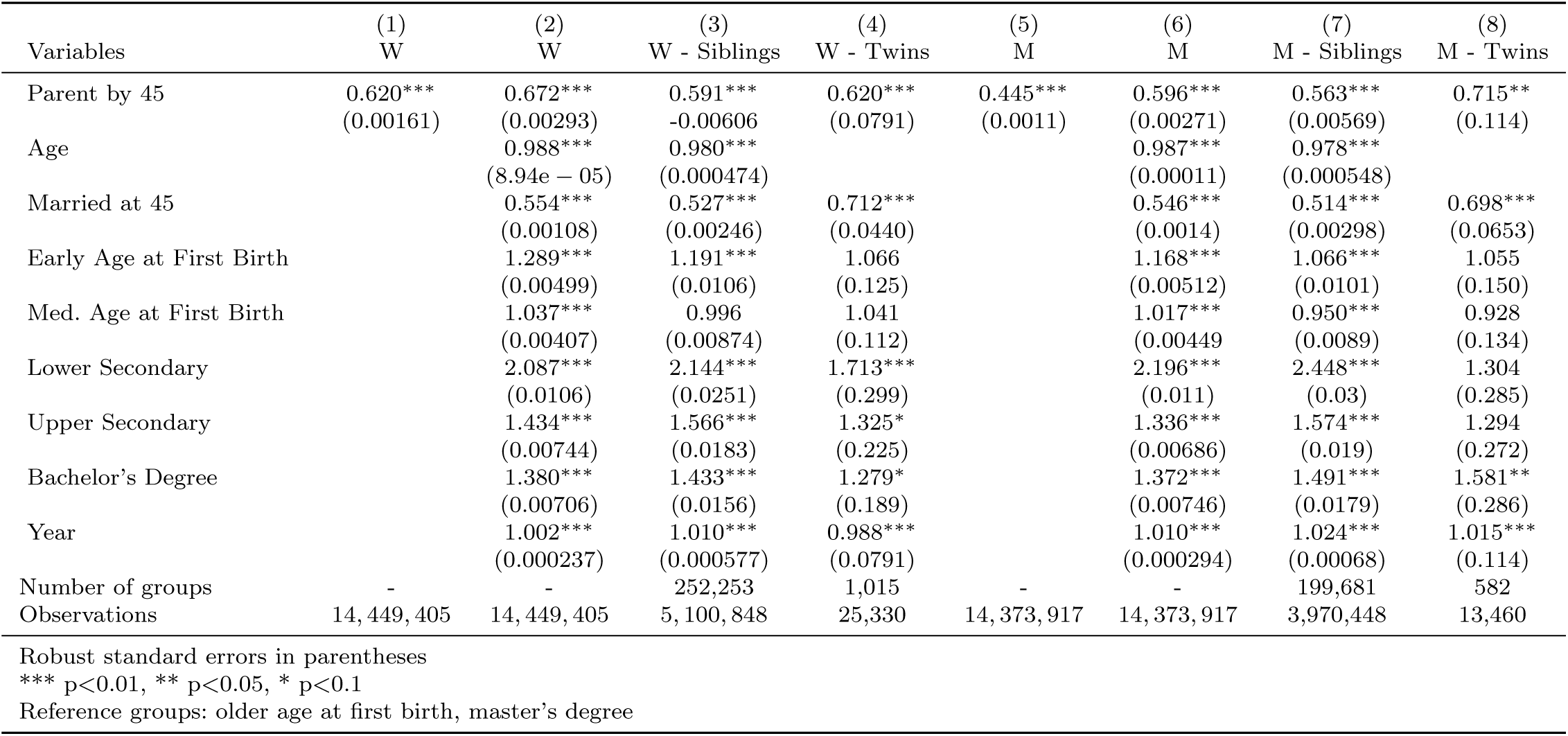
Mental Disorders (ORs)

**Table A3.**
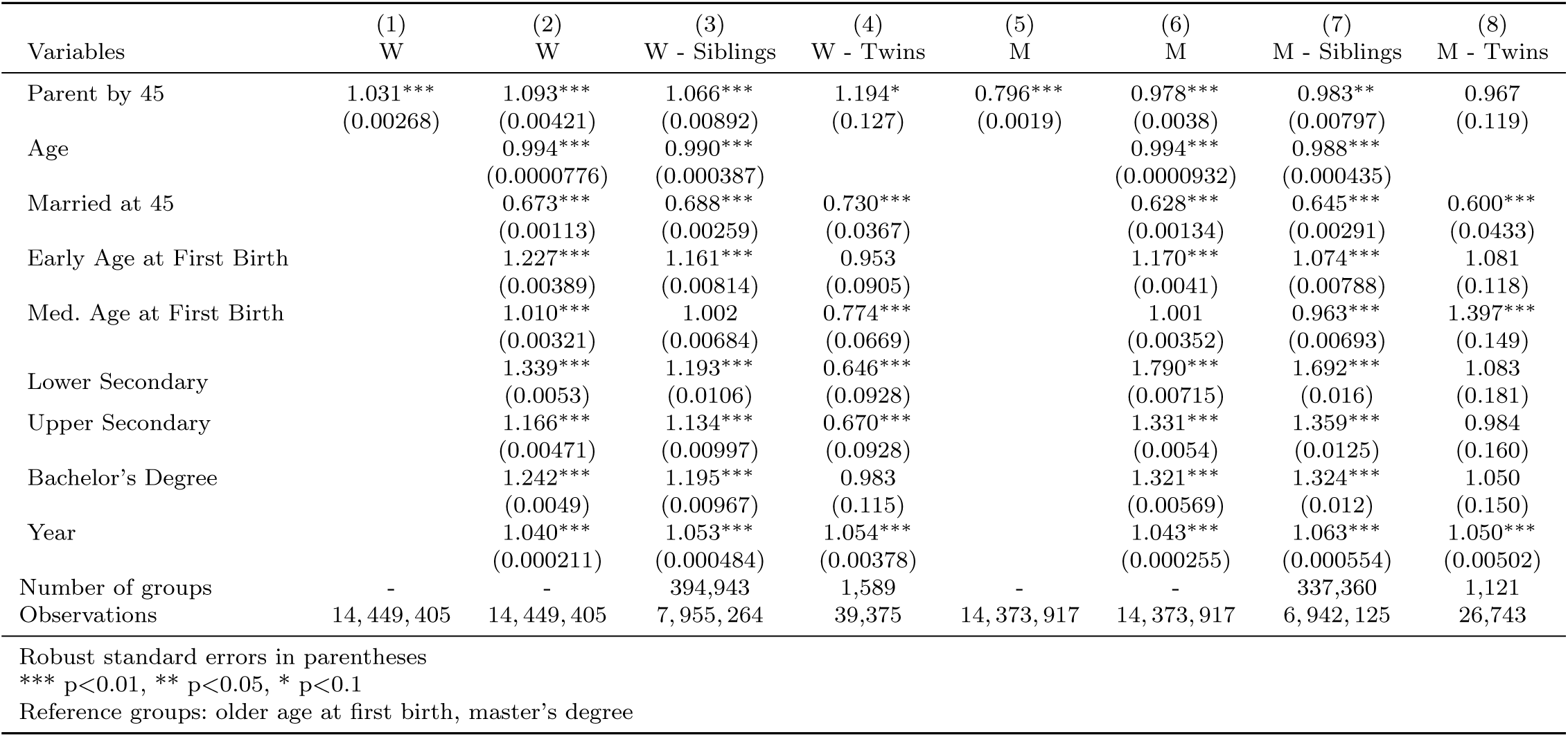
Psychological Symptoms/Complaints (ORs)

